# Convalescent Plasma and Improved Survival in Patients with Hematologic Malignancies and COVID-19

**DOI:** 10.1101/2021.02.05.21250953

**Authors:** Michael A. Thompson, Jeffrey P. Henderson, Pankil K. Shah, Samuel M. Rubinstein, Michael J. Joyner, Toni K. Choueiri, Daniel B. Flora, Elizabeth A. Griffiths, Anthony P. Gulati, Clara Hwang, Vadim S. Koshkin, Esperanza B. Papadopoulos, Elizabeth V. Robilotti, Christopher T. Su, Elizabeth M. Wulff-Burchfield, Zhuoer Xie, Peter Paul Yu, Sanjay Mishra, Jonathon W. Senefeld, Dimpy P. Shah, Jeremy L. Warner, on behalf of the COVID-19 and Cancer Consortium

## Abstract

Convalescent plasma may benefit immunocompromised individuals with COVID-19, including those with hematologic malignancy. We evaluated the association of convalescent plasma treatment with 30-day mortality in hospitalized adults with hematologic malignancy and COVID-19 from a multi-institutional cohort. 143 treated patients were compared to 823 untreated controls. After adjustment for potential confounding factors, convalescent plasma treatment was associated with improved 30-day mortality (hazard ratio, 0.60; 95% CI, 0.37-0.97). This association remained significant after propensity-score matching (hazard ratio, 0.52; 95% CI, 0.29-0.92). These findings suggest a potential survival benefit in the administration of convalescent plasma to patients with hematologic malignancy and COVID-19.

## BACKGROUND

Since initial reports in late 2019, SARS Coronavirus-2 (SARS-CoV-2) has infected over 100 million people worldwide and caused over 2 million deaths by early 2021.^1^ To date, data guiding COVID-19 therapies have largely arisen from large scale studies of healthy adults. Patients with hematologic malignancy represent a distinctive subset of COVID-19 patients due to immune deficits associated with both the diseases themselves and their treatments. Hematologic malignancy has been consistently associated with increased COVID-19 mortality and other complications.^2–4^

Antibody-based immunity is an important correlate of SARS-CoV-2 recovery and vaccine-associated prevention. Hematologic malignancies are associated with defects in humoral and cellular immunity that may contribute to adverse COVID-19 outcomes. Impaired antibody function is a well described complication of plasma cell neoplasms, chronic lymphocytic leukemia, and other lymphoid malignancies. Treatment of hematologic malignancies often exacerbate these immune defects; for example, rituximab targets the pan-B cell marker, CD20, and is highly effective therapy for B-cell malignancies. However, B-cell depletion can cause lymphopenia and hypogammaglobulinemia, and is associated with more severe COVID-19.^5^ Lymphopenia is known to be associated with more severe COVID-19.^6^

Antibody therapy using COVID-19 convalescent plasma was associated with a therapeutic benefit in a general patient population^7^ and older patients^8^ when high titer units were administered early in the course of disease. A negative prospective randomized trial included only 4 patients with hematologic malignancy in the convalescent plasma group.^9^ In immunodeficient patients, case reports have noted exceptional improvements in clinical status following convalescent plasma therapy, even following relatively late infusion.^10^ Given the absence of definitive prospective trial data in patients with hematologic malignancy, we conducted a retrospective cohort study to evaluate the hypothesis that convalescent plasma can correct defects in humoral deficiency and improve outcomes.

## METHODS

### Setting

The COVID-19 and Cancer Consortium (CCC19) is an international consortium aimed at understanding the clinical impact of COVID-19 on patients with cancer through an IRB-exempted comprehensive registry (NCT04354701). The methodology for CCC19 has been described and published previously.^11^ We analyzed data from hospitalized U.S. adults with a current or past diagnosis of hematologic malignancy diagnosed with confirmed or suspected SARS-CoV-2 infection in 2020 and reported from March 17^th^, 2020 to January 21^st^, 2021 (full list of contributors is in the **Supplementary Appendix**). Treatment exposure was defined as receiving convalescent plasma at any time during the COVID-19 illness. The exclusion criteria were: incomplete follow-up resulting in unknown death status, unknown or missing convalescent plasma exposure, age <18 years, mild COVID-19 not requiring hospitalization, and non-U.S. residence. The following data elements were obtained: age; sex; race and ethnic group; smoking status; comorbidities; the first recorded absolute lymphocyte count; type of hematologic malignancy; cancer status at COVID-19 diagnosis; Eastern Cooperative Oncology Group (ECOG) performance status prior to COVID-19; receipt and timing of anti-cancer treatment; baseline COVID-19 severity; level of care required; other anti-COVID-19 therapies (i.e., corticosteroids, remdesivir, tocilizumab, hydroxychloroquine); and U.S. census region of patient’s residence. The full data dictionary is provided in the **Supplementary Appendix**.

### Statistical Analysis

We calculated bivariate frequencies to examine the associations among the baseline characteristics and receipt of convalescent plasma. The primary endpoint was death within 30 days of COVID-19 diagnosis. Living patients had their data censored at 30 days from diagnosis. Crude and adjusted hazards ratios and 95% CI to estimate the association between convalescent plasma use and 30-day all-cause mortality were calculated using Cox proportional-hazards regression models. The primary analysis used propensity-score matching to help account for the nonrandomized treatment administration of convalescent plasma.^12^ Individual propensities for receipt of convalescent plasma treatment were estimated using a multivariable probit regression model with baseline covariate adjustment using covariates that were determined *a priori* based on published literature and clinical importance: age, sex, race/ethnicity, hematologic malignancy type, cancer status, cancer treatment timing, ECOG performance status, obesity, presence of type 2 diabetes mellitus, hypertension, renal comorbidities, pulmonary comorbidities, receipt of cytotoxic chemotherapy within 3 months of COVID-19 diagnosis, and trimester of diagnosis [January to April 2020; May to August 2020; September to December 2020]). For matching, the nearest-neighbor method with a 1:1 ratio (treated units:control units) and 0.2 standard deviation of the distance measure was applied to estimate the average treatment effect.^13^ Marginal hazard ratio along with 95% CI based on cluster-robust standard errors are reported. Kaplan-Meier survival curves were generated to compare survival probabilities using log-rank and stratified log-rank tests between convalescent plasma recipients and non-recipients for unmatched and matched samples, respectively. We conducted several sensitivity analyses to explore the robustness of the findings for the primary hypothesis against the model specifications, such as varying the caliper size by +/-0.1 and changing the matching order from the default maximum distance first to random order with different seeds. Exploratory subgroup analyses were conducted to determine whether patients with more severe illness (intensive care unit admission and/or mechanical ventilation) had differential outcome by convalescent plasma exposure.

We interpreted findings based on the 95% CI for the estimated measures of association. Reported P values are two-sided. Statistical analyses were performed using R software, version 4.0.3 (with packages MatchIt and Survival).

## RESULTS

As of January 21^st^, 2021, the CCC19 registry contained 8209 case reports with complete baseline information. 1761 (21.5%) patients had a primary or secondary hematologic malignancy, with lymphoid malignancies being the most common. After applying eligibility criteria (**Supplemental Figure S1**), there were n=966 patients, of whom 143 (14.8%) received convalescent plasma treatment (**Supplemental Figure S2**). Patient characteristics are noted in **Table 1**. Median age was 67 (interquartile range 58 to 77). In the unmatched sample, convalescent plasma recipients were slightly younger and more likely to be male. A lower proportion of convalescent plasma recipients had pulmonary comorbidities and ECOG performance status of 2 or more compared to the unexposed group. Convalescent plasma recipients were also more likely to be treated with corticosteroids, tocilizumab, and/or remdesivir, and less likely with hydroxychloroquine. Overall, 512 (53%) patients had received systemic anticancer treatment within 3 months of COVID-19 diagnosis, with targeted therapies (monoclonal antibodies, small molecule inhibitors, and/or immunomodulators) being the most commonly received treatments. 115 (22%) of those treated received an anti-CD20 antibody-containing regimen. Overall, 489 (58%) of 845 patients with absolute lymphocyte count available had lymphopenia (<1.5 x 10^9^ cells/liter) at presentation; this proportion increased to 79% in patients who had received anti-CD20 antibodies. Propensity-score matching was successful, with good balance achieved between the exposed and non-exposed groups (**Supplement Figures S3-S5**). Convalescent plasma recipients were more likely to require aggressive care (intensive care unit admission and/or mechanical ventilation).

**Table 1:**
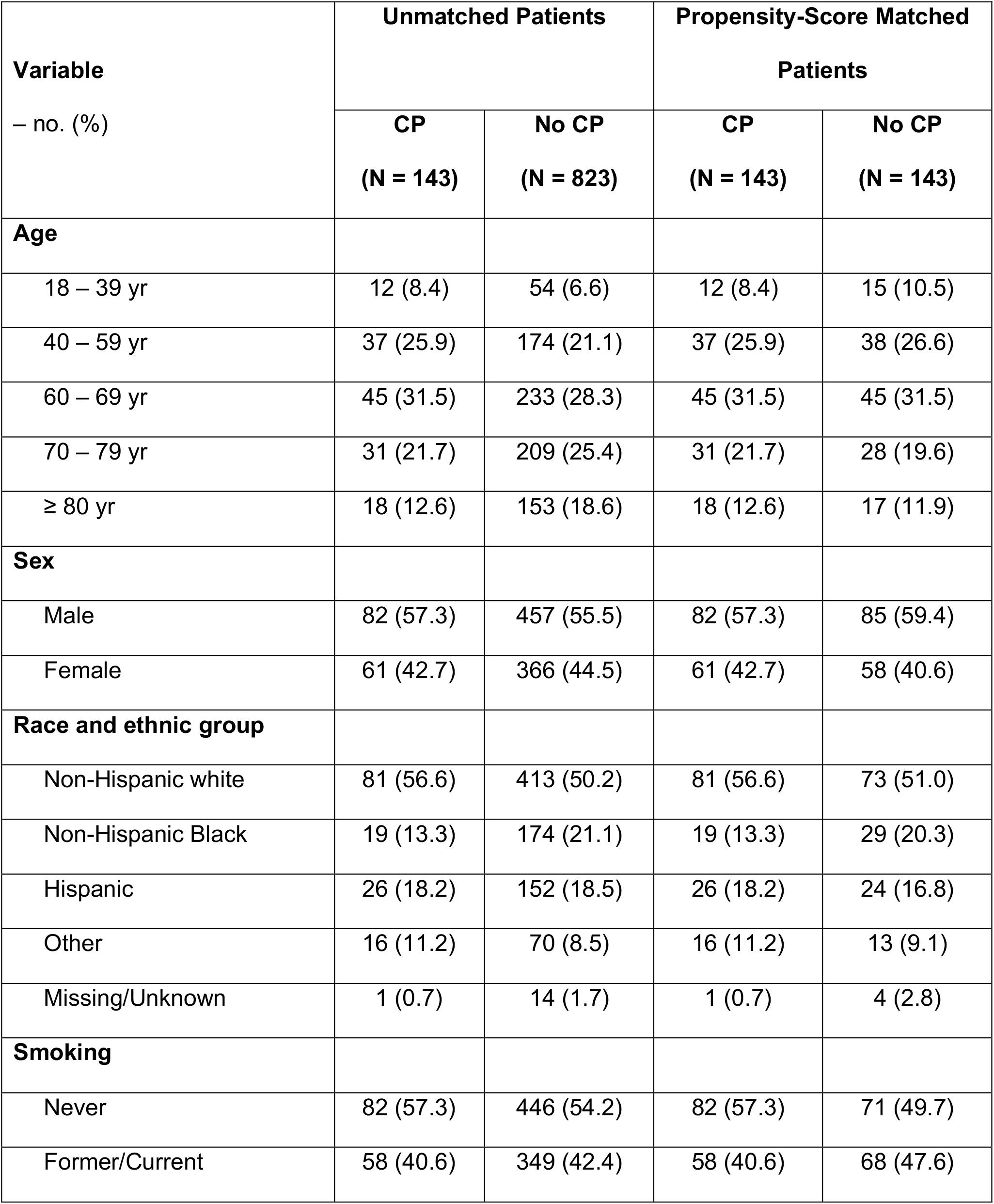
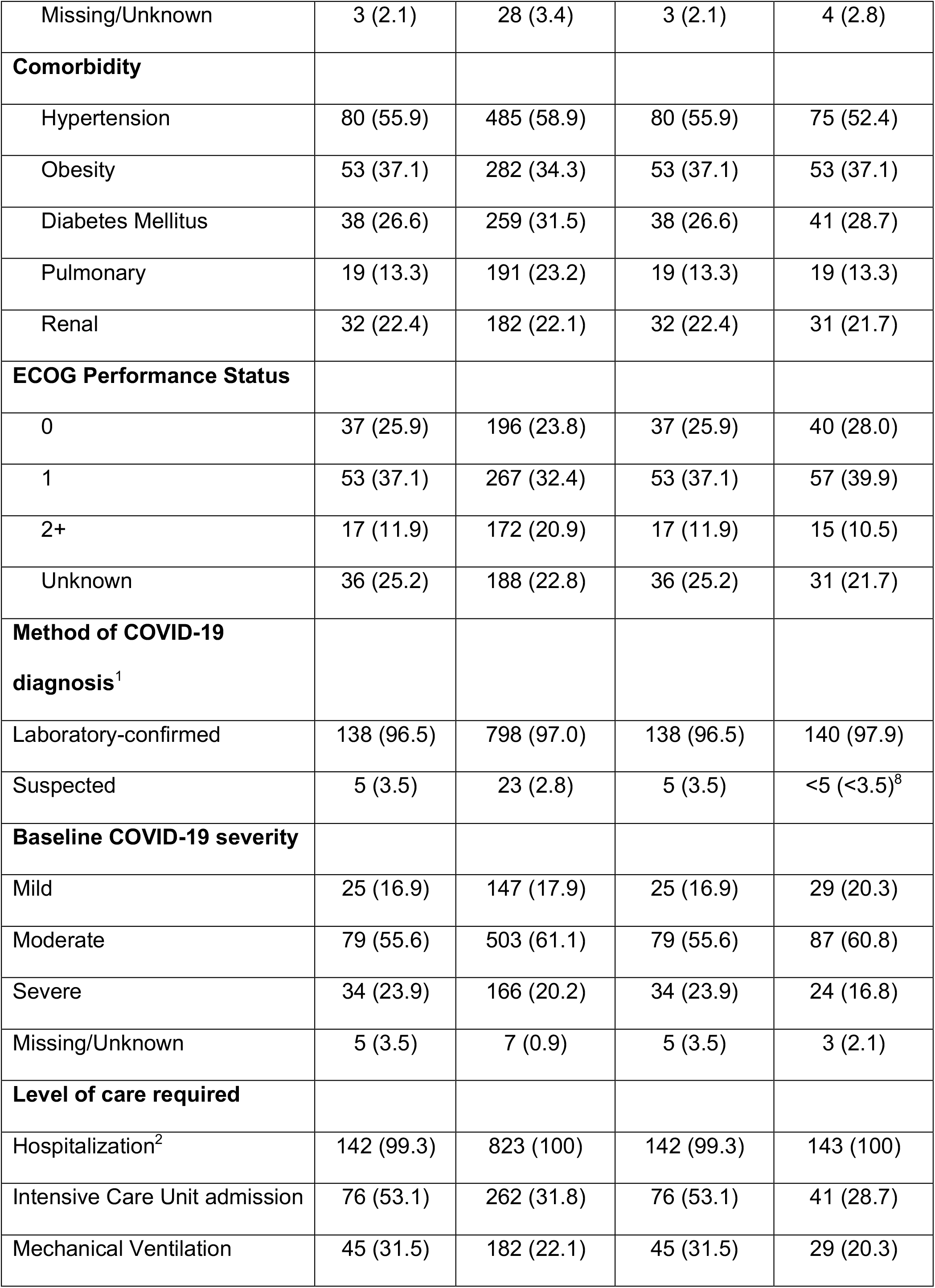
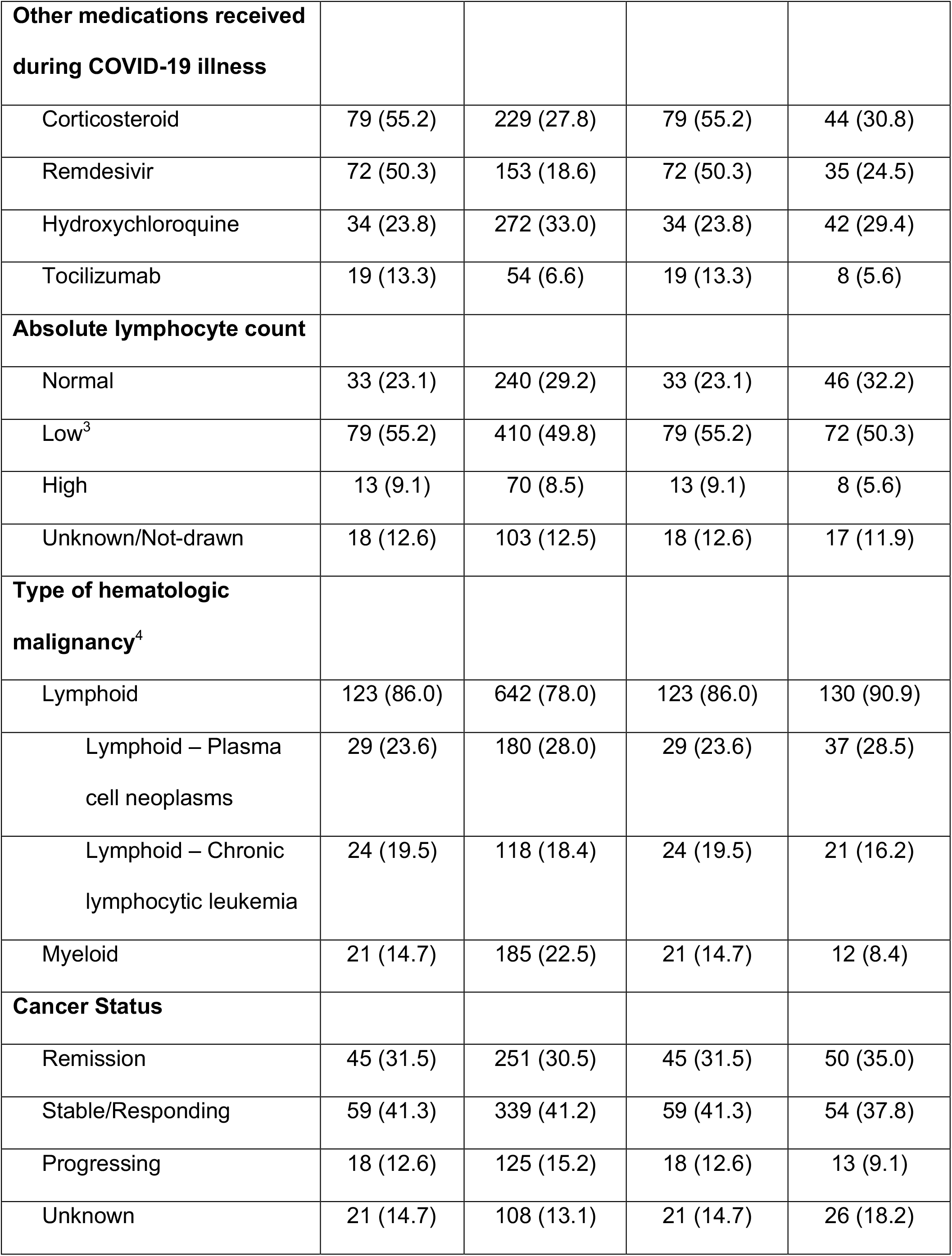
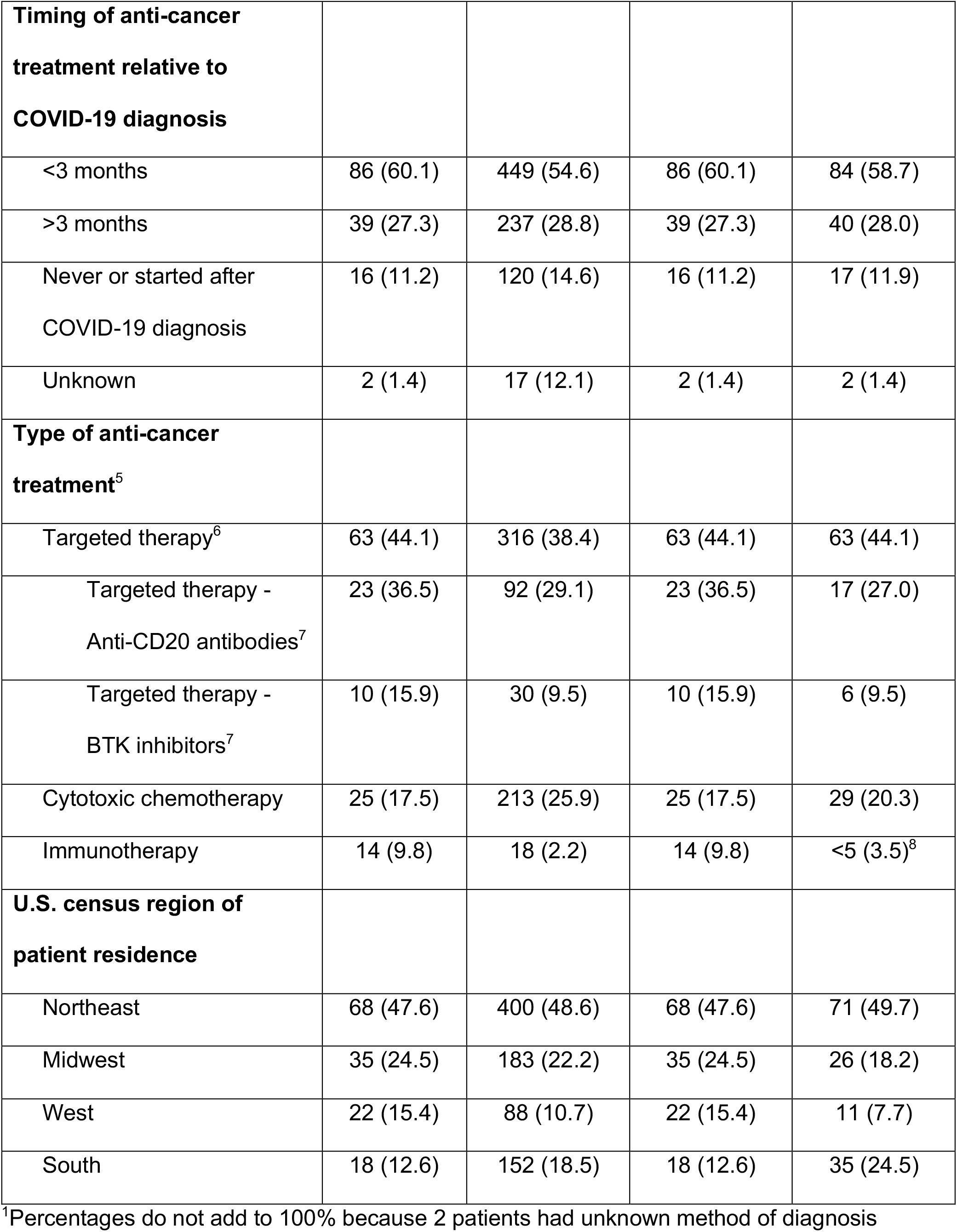
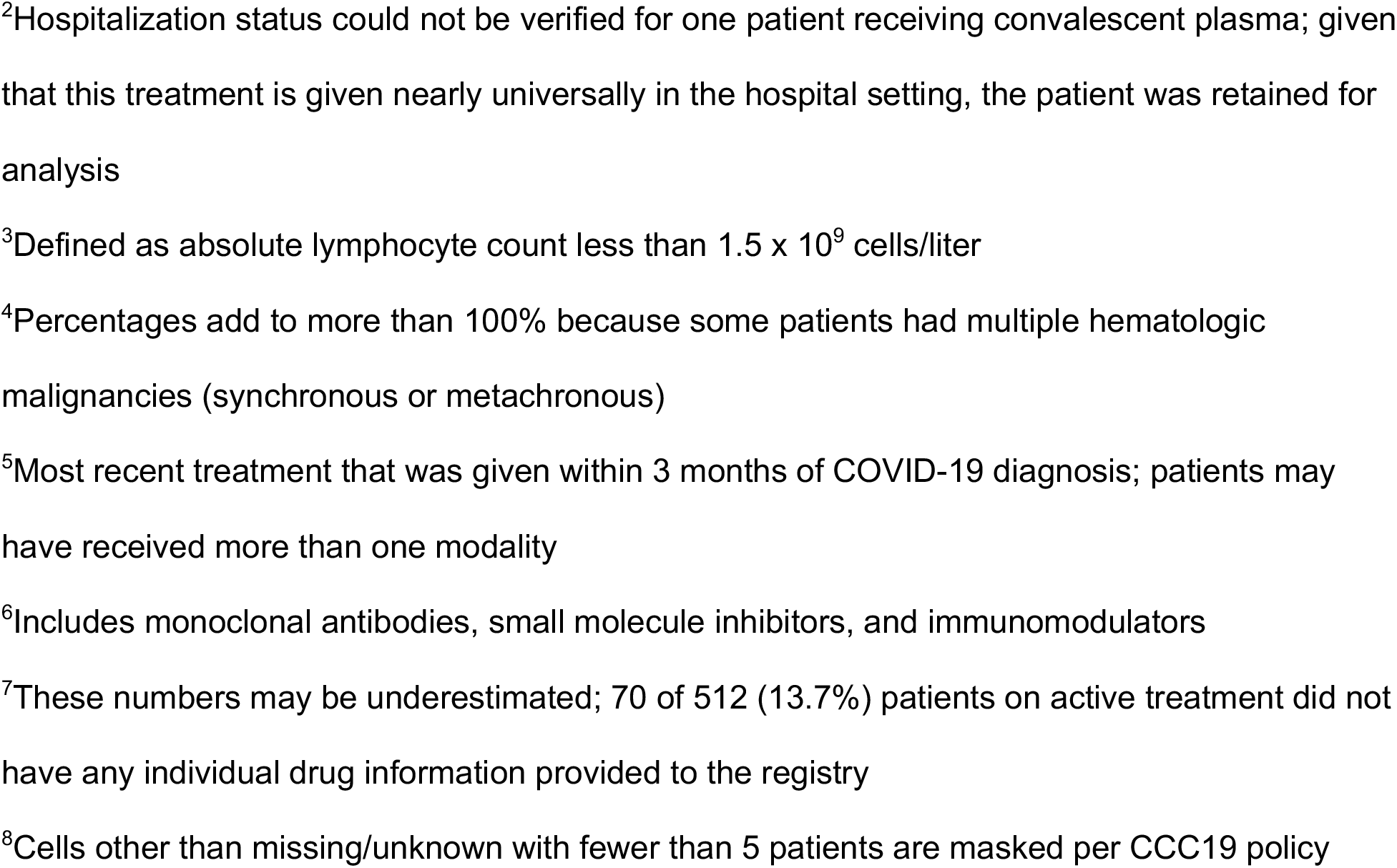
Characteristics of Patients Receiving or Not Receiving Convalescent Plasma before and after Propensity-Score Matching. CP: COVID-19 convalescent plasma

| **Variable**<br>– no. (%) | **Unmatched Patients** |  | **Propensity-Score Matched Patients** |  |
| --- | --- | --- | --- | --- |
|  | **CP**<br>**(N = 143)** | **No CP**<br>**(N = 823)** | **CP**<br>**(N = 143)** | **No CP**<br>**(N = 143)** |
| **Age** |  |  |  |  |
| 18 – 39 yr | 12 (8.4) | 54 (6.6) | 12 (8.4) | 15 (10.5) |
| 40 – 59 yr | 37 (25.9) | 174 (21.1) | 37 (25.9) | 38 (26.6) |
| 60 – 69 yr | 45 (31.5) | 233 (28.3) | 45 (31.5) | 45 (31.5) |
| 70 – 79 yr | 31 (21.7) | 209 (25.4) | 31 (21.7) | 28 (19.6) |
| ≥ 80 yr | 18 (12.6) | 153 (18.6) | 18 (12.6) | 17 (11.9) |
| **Sex** |  |  |  |  |
| Male | 82 (57.3) | 457 (55.5) | 82 (57.3) | 85 (59.4) |
| Female | 61 (42.7) | 366 (44.5) | 61 (42.7) | 58 (40.6) |
| **Race and ethnic group** |  |  |  |  |
| Non-Hispanic white | 81 (56.6) | 413 (50.2) | 81 (56.6) | 73 (51.0) |
| Non-Hispanic Black | 19 (13.3) | 174 (21.1) | 19 (13.3) | 29 (20.3) |
| Hispanic | 26 (18.2) | 152 (18.5) | 26 (18.2) | 24 (16.8) |
| Other | 16 (11.2) | 70 (8.5) | 16 (11.2) | 13 (9.1) |
| Missing/Unknown | 1 (0.7) | 14 (1.7) | 1 (0.7) | 4 (2.8) |
| **Smoking** |  |  |  |  |
| Never | 82 (57.3) | 446 (54.2) | 82 (57.3) | 71 (49.7) |
| Former/Current | 58 (40.6) | 349 (42.4) | 58 (40.6) | 68 (47.6) |
| Missing/Unknown | 3 (2.1) | 28 (3.4) | 3 (2.1) | 4 (2.8) |
| **Comorbidity** |  |  |  |  |
| Hypertension | 80 (55.9) | 485 (58.9) | 80 (55.9) | 75 (52.4) |
| Obesity | 53 (37.1) | 282 (34.3) | 53 (37.1) | 53 (37.1) |
| Diabetes Mellitus | 38 (26.6) | 259 (31.5) | 38 (26.6) | 41 (28.7) |
| Pulmonary | 19 (13.3) | 191 (23.2) | 19 (13.3) | 19 (13.3) |
| Renal | 32 (22.4) | 182 (22.1) | 32 (22.4) | 31 (21.7) |
| **ECOG Performance Status** |  |  |  |  |
| 0 | 37 (25.9) | 196 (23.8) | 37 (25.9) | 40 (28.0) |
| 1 | 53 (37.1) | 267 (32.4) | 53 (37.1) | 57 (39.9) |
| 2+ | 17 (11.9) | 172 (20.9) | 17 (11.9) | 15 (10.5) |
| Unknown | 36 (25.2) | 188 (22.8) | 36 (25.2) | 31 (21.7) |
| **Method of COVID-19 diagnosis<sup>1</sup>** |  |  |  |  |
| Laboratory-confirmed | 138 (96.5) | 798 (97.0) | 138 (96.5) | 140 (97.9) |
| Suspected | 5 (3.5) | 23 (2.8) | 5 (3.5) | <5 (<3.5)<sup>8</sup> |
| **Baseline COVID-19 severity** |  |  |  |  |
| Mild | 25 (16.9) | 147 (17.9) | 25 (16.9) | 29 (20.3) |
| Moderate | 79 (55.6) | 503 (61.1) | 79 (55.6) | 87 (60.8) |
| Severe | 34 (23.9) | 166 (20.2) | 34 (23.9) | 24 (16.8) |
| Missing/Unknown | 5 (3.5) | 7 (0.9) | 5 (3.5) | 3 (2.1) |
| **Level of care required** |  |  |  |  |
| Hospitalization<sup>2</sup> | 142 (99.3) | 823 (100) | 142 (99.3) | 143 (100) |
| Intensive Care Unit admission | 76 (53.1) | 262 (31.8) | 76 (53.1) | 41 (28.7) |
| Mechanical Ventilation | 45 (31.5) | 182 (22.1) | 45 (31.5) | 29 (20.3) |
| **Other medications received during COVID-19 illness** |  |  |  |  |
| Corticosteroid | 79 (55.2) | 229 (27.8) | 79 (55.2) | 44 (30.8) |
| Remdesivir | 72 (50.3) | 153 (18.6) | 72 (50.3) | 35 (24.5) |
| Hydroxychloroquine | 34 (23.8) | 272 (33.0) | 34 (23.8) | 42 (29.4) |
| Tocilizumab | 19 (13.3) | 54 (6.6) | 19 (13.3) | 8 (5.6) |
| **Absolute lymphocyte count** |  |  |  |  |
| Normal | 33 (23.1) | 240 (29.2) | 33 (23.1) | 46 (32.2) |
| Low<sup>3</sup> | 79 (55.2) | 410 (49.8) | 79 (55.2) | 72 (50.3) |
| High | 13 (9.1) | 70 (8.5) | 13 (9.1) | 8 (5.6) |
| Unknown/Not-drawn | 18 (12.6) | 103 (12.5) | 18 (12.6) | 17 (11.9) |
| **Type of hematologic malignancy<sup>4</sup>** |  |  |  |  |
| Lymphoid | 123 (86.0) | 642 (78.0) | 123 (86.0) | 130 (90.9) |
| Lymphoid – Plasma cell neoplasms | 29 (23.6) | 180 (28.0) | 29 (23.6) | 37 (28.5) |
| Lymphoid – Chronic lymphocytic leukemia | 24 (19.5) | 118 (18.4) | 24 (19.5) | 21 (16.2) |
| Myeloid | 21 (14.7) | 185 (22.5) | 21 (14.7) | 12 (8.4) |
| **Cancer Status** |  |  |  |  |
| Remission | 45 (31.5) | 251 (30.5) | 45 (31.5) | 50 (35.0) |
| Stable/Responding | 59 (41.3) | 339 (41.2) | 59 (41.3) | 54 (37.8) |
| Progressing | 18 (12.6) | 125 (15.2) | 18 (12.6) | 13 (9.1) |
| Unknown | 21 (14.7) | 108 (13.1) | 21 (14.7) | 26 (18.2) |
| **Timing of anti-cancer treatment relative to COVID-19 diagnosis** |  |  |  |  |
| <3 months | 86 (60.1) | 449 (54.6) | 86 (60.1) | 84 (58.7) |
| >3 months | 39 (27.3) | 237 (28.8) | 39 (27.3) | 40 (28.0) |
| Never or started after COVID-19 diagnosis | 16 (11.2) | 120 (14.6) | 16 (11.2) | 17 (11.9) |
| Unknown | 2 (1.4) | 17 (12.1) | 2 (1.4) | 2 (1.4) |
| **Type of anti-cancer treatment<sup>5</sup>** |  |  |  |  |
| Targeted therapy<sup>6</sup> | 63 (44.1) | 316 (38.4) | 63 (44.1) | 63 (44.1) |
| Targeted therapy - Anti-CD20 antibodies<sup>7</sup> | 23 (36.5) | 92 (29.1) | 23 (36.5) | 17 (27.0) |
| Targeted therapy - BTK inhibitors<sup>7</sup> | 10 (15.9) | 30 (9.5) | 10 (15.9) | 6 (9.5) |
| Cytotoxic chemotherapy | 25 (17.5) | 213 (25.9) | 25 (17.5) | 29 (20.3) |
| Immunotherapy | 14 (9.8) | 18 (2.2) | 14 (9.8) | <5 (3.5)<sup>8</sup> |
| **U.S. census region of patient residence** |  |  |  |  |
| Northeast | 68 (47.6) | 400 (48.6) | 68 (47.6) | 71 (49.7) |
| Midwest | 35 (24.5) | 183 (22.2) | 35 (24.5) | 26 (18.2) |
| West | 22 (15.4) | 88 (10.7) | 22 (15.4) | 11 (7.7) |
| South | 18 (12.6) | 152 (18.5) | 18 (12.6) | 35 (24.5) |

<sup>1</sup>Percentages do not add to 100% because 2 patients had unknown method of diagnosis
<sup>2</sup>Hospitalization status could not be verified for one patient receiving convalescent plasma; given that this treatment is given nearly universally in the hospital setting, the patient was retained for analysis
<sup>3</sup>Defined as absolute lymphocyte count less than $1.5 \times 10^9$ cells/liter
<sup>4</sup>Percentages add to more than 100% because some patients had multiple hematologic malignancies (synchronous or metachronous)
<sup>5</sup>Most recent treatment that was given within 3 months of COVID-19 diagnosis; patients may have received more than one modality
<sup>6</sup>Includes monoclonal antibodies, small molecule inhibitors, and immunomodulators
<sup>7</sup>These numbers may be underestimated; 70 of 512 (13.7%) patients on active treatment did not have any individual drug information provided to the registry
<sup>8</sup>Cells other than missing/unknown with fewer than 5 patients are masked per CCC19 policy

With a median follow-up period of 30 days (interquartile range 21 to 90 days), there were 223 (23.1%) deaths within 30 days of COVID-19 diagnosis (**Table 2**). The crude hazard rate for mortality was significantly lower in convalescent plasma recipients (19 out of 143; 13.3%), compared to non-recipients (204 out of 823; 24.8%). This difference was statistically significant after adjustment in the overall comparison (hazard ratio, 0.60; 95% CI, 0.37 to 0.97; P=0.033) and the propensity-score matched comparison (hazard ratio, 0.52; 95% CI, 0.29 to 0.92; P=0.025) (**Table 2**). Non-recipients had significantly lower overall survival than those who received convalescent plasma in the unmatched and matched samples (**Figure 1**). Multiple additional sensitivity analyses, including analyses that used different caliper sizes for matching and analyses with randomized matching orders, showed similar results. Among the 338 patients admitted to the intensive care unit, the crude hazard rate for mortality was significantly lower in convalescent plasma recipients compared to non-recipients in the overall comparison (adjusted hazard ratio, 0.30; 95% CI, 0.16 to 0.56) and the propensity-score matched comparison (hazard ratio, 0.40; 95% CI, 0.20 to 0.80). Among the 227 patients requiring mechanical ventilation, the crude hazard rate for mortality was significantly lower in convalescent plasma recipients compared to non-recipients in the overall comparison (hazard ratio, 0.23; 95% CI, 0.10 to 0.50) and the propensity-score matched comparison (hazard ratio, 0.32; 95% CI, 0.14 to 0.72) (**Table 2; Supplemental Figure S6**).

**Table 2:**
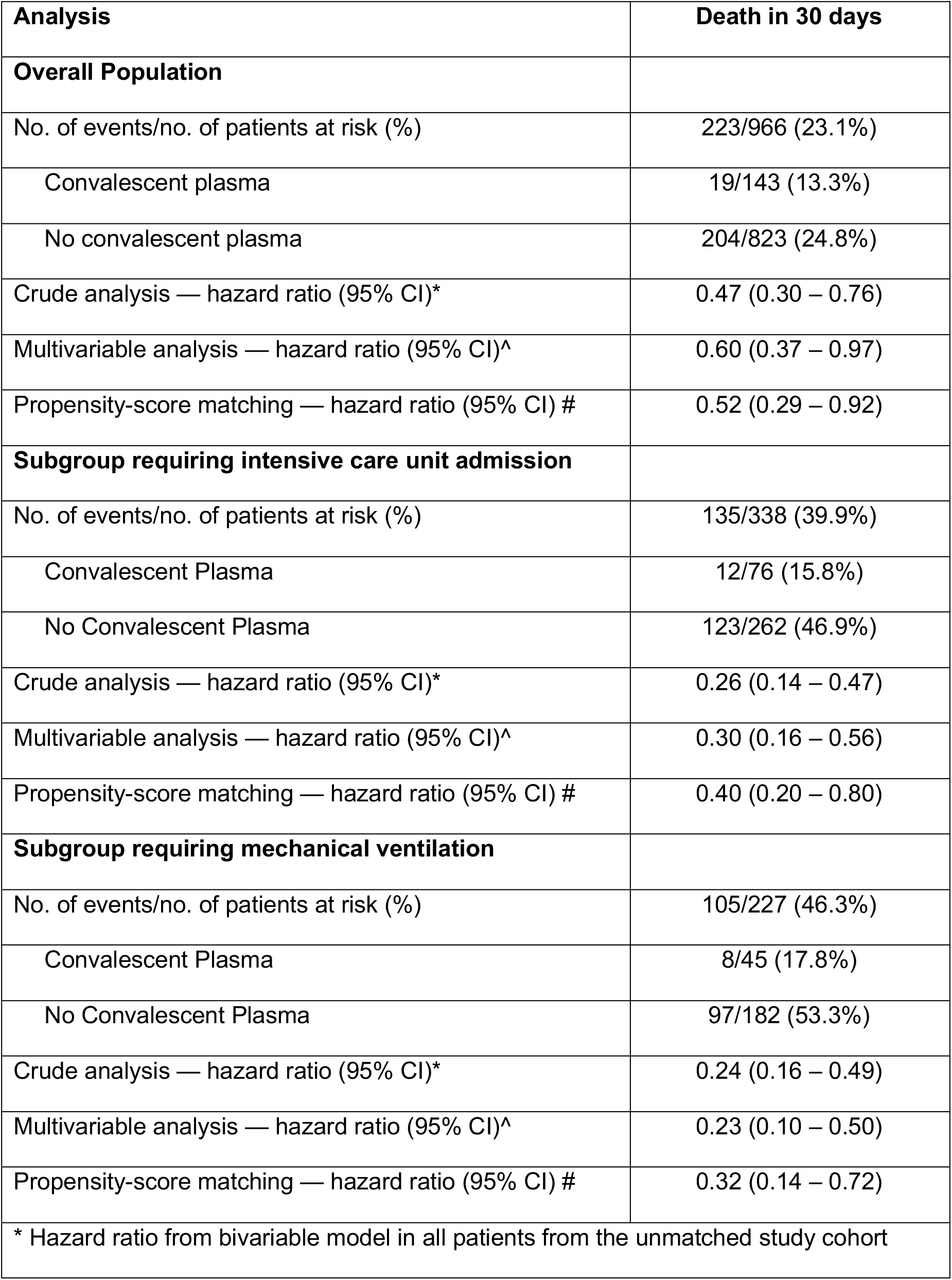
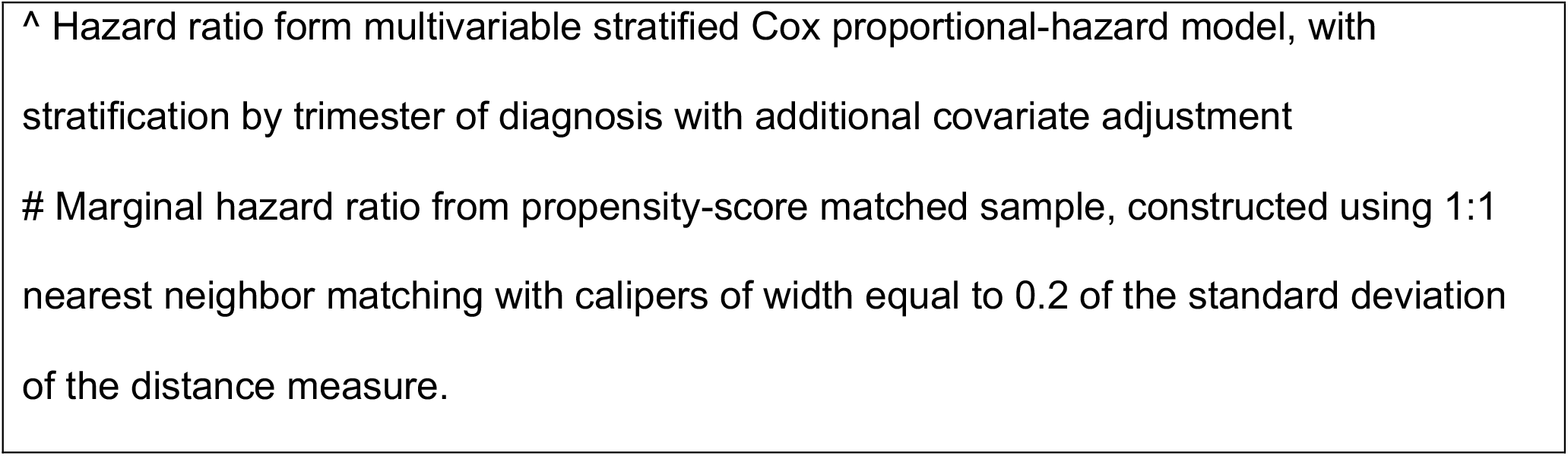
Association between Convalescent Plasma use and Death in the Crude Analysis, Multivariable Analysis, and Propensity-Score Analyses.

| **Analysis** | **Death in 30 days** |
| --- | --- |
| **Overall Population** |  |
| No. of events/no. of patients at risk (%) | 223/966 (23.1%) |
| Convalescent plasma | 19/143 (13.3%) |
| No convalescent plasma | 204/823 (24.8%) |
| Crude analysis — hazard ratio (95% CI)* | 0.47 (0.30 – 0.76) |
| Multivariable analysis — hazard ratio (95% CI)^ | 0.60 (0.37 – 0.97) |
| Propensity-score matching — hazard ratio (95% CI) # | 0.52 (0.29 – 0.92) |
| **Subgroup requiring intensive care unit admission** |  |
| No. of events/no. of patients at risk (%) | 135/338 (39.9%) |
| Convalescent Plasma | 12/76 (15.8%) |
| No Convalescent Plasma | 123/262 (46.9%) |
| Crude analysis — hazard ratio (95% CI)* | 0.26 (0.14 – 0.47) |
| Multivariable analysis — hazard ratio (95% CI)^ | 0.30 (0.16 – 0.56) |
| Propensity-score matching — hazard ratio (95% CI) # | 0.40 (0.20 – 0.80) |
| **Subgroup requiring mechanical ventilation** |  |
| No. of events/no. of patients at risk (%) | 105/227 (46.3%) |
| Convalescent Plasma | 8/45 (17.8%) |
| No Convalescent Plasma | 97/182 (53.3%) |
| Crude analysis — hazard ratio (95% CI)* | 0.24 (0.16 – 0.49) |
| Multivariable analysis — hazard ratio (95% CI)^ | 0.23 (0.10 – 0.50) |
| Propensity-score matching — hazard ratio (95% CI) # | 0.32 (0.14 – 0.72) |
| * Hazard ratio from bivariable model in all patients from the unmatched study cohort |  |

^ Hazard ratio from multivariable stratified Cox proportional-hazard model, with stratification by trimester of diagnosis with additional covariate adjustment

**Figure 1:**
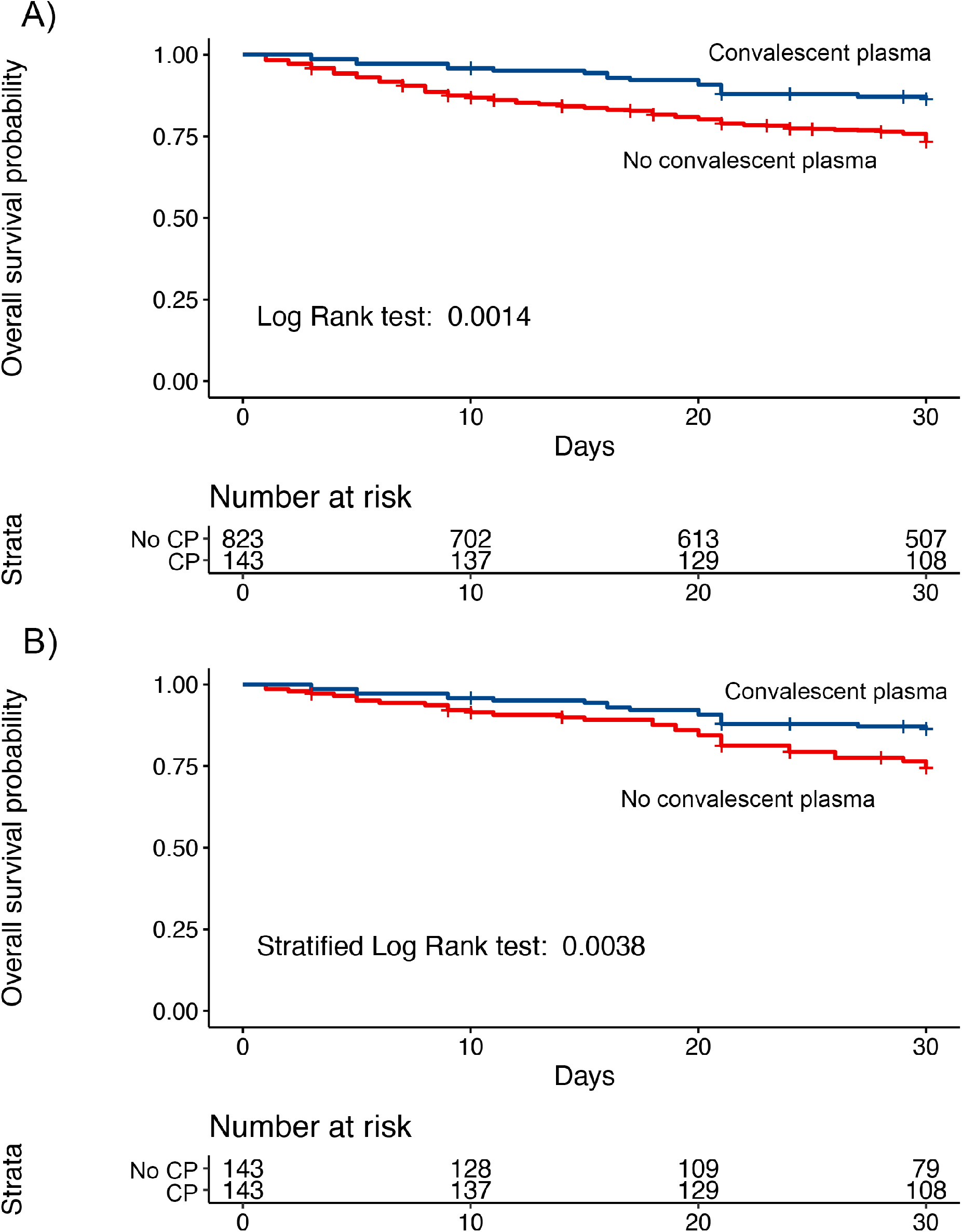
Primary Outcome in the A) Overall Population and B) Propensity-Score Matched Population.

## DISCUSSION

There is historical evidence demonstrating the efficacy of passive antibody therapy for infectious diseases when given early in the course of disease prior to the development of endogenous antibody responses, including in severe acute respiratory infections.^14–16^ On this basis, interventional trials of convalescent plasma in COVID-19 are ongoing; to our knowledge, only one of these, FALP-COVID (NCT04384588) is specifically recruiting patients with cancer. There is accumulating evidence supporting the efficacy of convalescent plasma in patients with primary or secondary immunodeficiency, including those subjected to profound immunosuppression in the setting of hematopoietic stem cell transplantation.^17,18^ Patients with hematologic malignancies may have immune deficiencies from patient factors (including age), disease factors, and treatment factors. For example, in a single center cohort of patients with chronic lymphocytic leukemia (a common B-cell hematologic malignancy) who had documented symptomatic COVID-19, 7 of 21 (33%) did not develop detectable anti-SARS-CoV-2 antibodies, notably lower than the 100% seroconversion rate observed in a non-cancer population.^19,20^ Several small studies have demonstrated improvement in clinical course after administration of convalescent to patients with cancer, primarily hematologic malignancy.^21–23^ Clinical improvement in COVID-19 symptoms within 48 hours of convalescent plasma transfusion was also reported in 16 of 17 patients with B-cell lymphopenia and prolonged COVID-19, 15 of whom had received anti-CD20 therapy in the 3-6 months prior to symptoms onset.^23^

Lymphopenia was common in our study population, especially in patients with recent anti-CD20 treatment, as would be expected. We are unable to ascertain rates of hypogammaglobulinemia, as this was not a routinely collected variable. The exact mechanism by which convalescent plasma may have mediated improved outcomes in the treated patients is likely multifactorial, and could include reduction in viral load via enhanced clearance,^23^ reduction in secondary bacterial and fungal infections, neutralization of inflammatory cytokines that may otherwise promote a hyperinflammatory immune phenotype,^24^ and temporizing until the native immune system generates additional humoral and cell-mediated responses in the recovery phase after myelosuppressive or lymphodepleting anticancer therapy.

Our study is the largest such series reported to date, to our knowledge. Due to the multi-institutional nature of the data with over 70 contributing institutions (**Supplemental Appendix**), these findings are unlikely to be the result of specific practice patterns at certain institutions. Variables collected through this effort, such as cancer status, prior cancer treatments, and ECOG performance status, are not readily available through automated electronic health record extractions or claims databases.

Limitations of this study include its retrospective nature, as well as unmeasured variables, such as the exact timing of convalescent plasma administration with respect to the date of COVID-19 diagnosis, the antibody titers/levels in the plasma that was administered, and whether repeat dosing was employed. Despite propensity matching, it is possible that residual confounding remains, and results should be interpreted with caution. For example, even after propensity matching, the convalescent plasma recipients received more corticosteroids and remdesivir. Although these agents have not been shown to have a clear survival benefit in cancer populations,^25^ it is possible that at least part of the observed protective effect of convalescent plasma could be due to concomitant medications, including fewer administrations of hydroxychloroquine. Convalescent plasma non-recipients may have received less aggressive care overall due to factors other than COVID-19, e.g., advanced states of cancer; this possibility is partially addressed through adjustment for cancer status. Differential access to convalescent plasma due to health system or socioeconomic factors cannot be excluded. It is possible that the findings in the first 30 days would not persist into later periods, which would require a more extended follow-up. Therefore, like any observational study, these findings should not be inferred for causality but rather be seen as contributing to the accumulating evidence regarding survival benefit with convalescent plasma in patients with COVID-19 illness. Prospective randomized trials evaluating convalescent plasma in patients with hematologic malignancy with attention to administration timing and consideration of repeated dosing are recommended.

In conclusion, convalescent plasma appears to confer a survival benefit in patients with hematologic malignancy and COVID-19. If this finding should hold up in prospective clinical trials, convalescent plasma would be, to our knowledge, the first COVID-19 intervention with a survival benefit in this high-risk population.

## Data Availability

The full CCC19 data dictionary and R code to generate the derived variables is publicly available on GitHub.
The data used for the analyses will be made available in aggregate form through a dashboard on the ccc19.org website, six months from the peer-reviewed publication of this research.

https://github.com/covidncancer/CCC19_dictionary

## ACKNOWLEDGMENTS

We thank all members of the CCC19 steering committee: Toni K. Choueiri, Dimitrios Farmakiotis, Petros Grivas, Gilberto de Lima Lopes Jr., Corrie A. Painter, Solange Peters, Brian I. Rini, Dimpy P. Shah, Michael A. Thompson, and Jeremy L. Warner, for their invaluable guidance of the CCC19 consortium. This project has been funded in whole or in part with Federal funds from the Department of Health and Human Services; Office of the Assistant Secretary for Preparedness and Response; Biomedical Advanced Research and Development Authority under Contract No. 75A50120C00096; National Cancer Institute (NCI) grants P30 CA008748, P30 CA046592, P30 CA054174, P30 CA068485, T32 CA236621, and U01 CA231840; National Center for Advancing Translational Sciences (NCATS) grant UL1 TR002377; Schwab Charitable Fund (Eric E Schmidt, Wendy Schmidt donors); United Health Group; National Basketball Association (NBA); Millennium Pharmaceuticals; Octapharma USA, Inc; the American Cancer Society and Hope Foundation for Cancer Research grant MRSG-16-152-01-CCE; the Longer Life Foundation: A RGA/Washington University Partnership; and the Mayo Clinic. REDCap is developed and supported by Vanderbilt Institute for Clinical and Translational Research grant support (UL1 TR000445 from NCATS/NIH).

## Disclaimer

The views and opinions expressed in this publication are those of the authors and do not reflect the official policy or position of the US Department of Health and Human services and its agencies including the Biomedical Research and Development Authority and the Food and Drug Administration, as well as any agency of the U.S. government. Assumptions made within and interpretations from the analysis are not reflective of the position of any U.S. government entity.

