## Supplemental Appendix for "Convalescent Plasma and Improved Survival in Patients with Hematologic Malignancies and COVID-19"

### Alphabetical list of participants by institution that contributed 1+ patients to the analysis

**Bolded = site PI/co-PIs)**; site co-Is are listed alphabetically by last name

**Balazs Halmos, MD**; **Amit Verma, MBBS**; Benjamin A. Gartrell, MD; Sanjay Goel, MBBS; Nitin Ohri, MD; R. Alejandro Sica, MD; Astha Thakkar, MD (Albert Einstein College of Medicine, Montefiore Medical Center, Bronx, NY, USA)

**Keith Stockerl-Goldstein, MD**; Omar Butt, MD, PhD; Jian L. Campian, MD, PhD; Mark A. Fiala, MSW; Ryan Monahan, MBA; Alice Y. Zhou, MD, PhD (Alvin J. Siteman Cancer Center at Washington University School of Medicine and Barnes-Jewish Hospital, St. Louis, MO, USA)

**Michael A. Thompson, MD, PhD, FASCO**; Pamela Bohachek, RN; Daniel Mundt, MD; Mitrianna Streckfuss, MPH; Eyob Tadesse, MD (Aurora Cancer Care, Advocate Aurora Health, Milwaukee, WI, USA)

**Philip E. Lammers, MD, MSCI** (Baptist Cancer Center, Memphis, TN, USA)

**Sanjay G. Revankar, MD, FIDSA** (The Barbara Ann Karmanos Cancer Institute at Wayne State University School of Medicine, Detroit, MI, USA)

**Orestis A. Panagiotou, MD, PhD**; Pamela C. Egan, MD; Dimitrios Farmakiotis, MD, FACP, FIDSA; Hina Khan, MD; Adam J. Olszewski, MD (Brown University and Lifespan Cancer Institute, Providence, RI, USA)

**Arturo Loaiza-Bonilla, MD, MSEd, FACP** (Cancer Treatment Centers of America, AZ/GA/IL/OK/PA, USA)

**Salvatore A. Del Prete, MD**; Anne H. Angevine, MD; Michael H. Bar, MD, FACP; Anthony P. Gulati, MD; K. M. Steve Lo, MD; Jamie Stratton, MD; Paul L. Weinstein, MD (Carl & Dorothy Bennett Cancer Center at Stamford Hospital, Stamford, CT, USA)

**Paolo Caimi, MD**; Jill S. Barnholtz-Sloan, PhD; Jorge A. Garcia, MD, FACP; John M. Nakayama, MD (Case Comprehensive Cancer Center at Case Western Reserve University/University Hospitals, Cleveland, OH, USA)

**Shilpa Gupta, MD; Nathan A. Pennell, MD, PhD, FASCO**; Manmeet S. Ahluwalia, MD, FACP; Scott J. Dawsey, MD; Christopher A. Lemmon, MD; Amanda Nizam, MD (Cleveland Clinic, Cleveland, OH, USA)

**Claire Hoppenot, MD; Ang Li, MD, MS** (Dan L Duncan Comprehensive Cancer Center at Baylor College of Medicine, Houston, TX, USA)

**Toni K. Choueiri, MD**; Ziad Bakouny, MD, MSc; Gabrielle Bouchard, BS; Fiona J. Busser, BA; Jean M. Connors, MD; Catherine Curran, BA; George D. Demetri, MD, FASCO; Antonio Giordano, MD, PhD; Kaitlin Kelleher, BA; Anju Nohria, MD; Andrew Schmidt, MD; Grace Shaw, BA; Eli Van Allen, MD; Pier Vitale Nuzzo, MD, PhD; Wenxin (Vincent) Xu, MD; Rebecca L. Zon, MD (Dana-Farber Cancer Institute, Boston, MA, USA)

**Tian Zhang, MD, MHS**; Susan Halabi, PhD, FASCO (Duke Cancer Institute at Duke University Medical Center, Durham, NC, USA)

**Gary H. Lyman, MD, MPH, FASCO, FRCP**; Jerome J. Graber MD, MPH; Petros Grivas, MD, PhD; Ali Raza Khaki, MD; Elizabeth T. Loggers, MD, PhD; Ryan C. Lynch, MD; Elizabeth S. Nakasone, MD, PhD; Michael T. Schweizer, MD; Lisa Tachiki, MD; Shaveta Vinayak, MD, MS; Michael J. Wagner, MD; Albert Yeh, MD (Fred Hutchinson Cancer Research Center/University of Washington/Seattle Cancer Care Alliance, Seattle, WA, USA)

**Na Tosha N. Gatson, MD, PhD** (Geisinger Health System, PA, USA)

**Sharad Goyal, MD; Minh-Phuong Huynh-Le, MD, MAS** (George Washington University, Washington, DC, USA)

**Lori J. Rosenstein, MD** (Gundersen Health System, WI, USA)

**Peter Paul Yu, MD, FACP, FASCO**; Jessica M. Clement, MD; Ahmad Daher, MD; Mark Dailey, MD; Rawad Elias, MD; Asha Jayaraj, MD; Emily Hsu, MD; Alvaro G. Menendez, MD; Joerg Rathmann, MD; Oscar Serrano, MD (Hartford HealthCare Cancer Institute, Hartford, CT, USA)

**Clara Hwang, MD**; Shirish M. Gadgeel, MD (Henry Ford Cancer Institute, Henry Ford Hospital, Detroit, MI, USA)

**Jessica E. Hawley, MD; Dawn Hershman, MD, MS, FASCO**; Melissa K. Accordino, MD, MS; Divaya Bhutani, MD; Gary K. Schwartz, MD (Herbert Irving Comprehensive Cancer Center at Columbia University, New York, NY, USA)

**Daniel Y. Reuben, MD, MS**; Sarah Mushtaq, MD (Hollings Cancer Center at the Medical University of South Carolina, Charleston, SC, USA)

**Eric H. Bernicker, MD** (Houston Methodist Cancer Center, Houston, TX, USA)

**John Deeken, MD**; Danielle Shafer, DO (Inova Schar Cancer Institute, Fairfax, VA, USA)

**Mark A. Lewis, MD**; **Terence D. Rhodes, MD, PhD**; David M. Gill, MD; Clarke A. Low; MD (Intermountain Health Care, Salt Lake City, UT, USA)

**Sarah Nagle, MD**; Shannon McWeeney, PhD; Eneida R. Nemecek, MD, MS, MBA (Knight Cancer Institute at Oregon Health and Science University, Portland, OR, USA)

**Howard A. Zaren, MD, FACS**, Stephanie J. Smith, RN, MSN, OCN (Lewis Cancer & Research Pavilion @ St. Joseph's/Candler, Savannah, GA, USA)

**Gayathri Nagaraj, MD**; Mojtaba Akhtari, MD; Eric Lau, DO; Mark E. Reeves, MD (Loma Linda University Cancer Center, Loma Linda, CA, USA)

**Stephanie Berg, DO**; Destry Elms, MD (Loyola University Medical Center, Maywood, IL, USA)

**Alicia K. Morgans, MD, MPH; Firas H. Wehbe, MD, PhD**; Jessica Altman, MD; Michael Gurley, BA; Mary F. Mulcahy, MD (Lurie Cancer Center at Northwestern University, Chicago, IL, USA)

**Eric B. Durbin, DrPH, MS** (Markey Cancer Center at the University of Kentucky, Lexington, KY, USA)

**Amit A. Kulkarni, MD**; Heather H. Nelson, PhD, MPH; Surbhi Shah, MD (Masonic Cancer Center at the University of Minnesota, Minneapolis, MN, USA)

**Rachel P. Rosovsky, MD, MPH; Kerry Reynolds, MD**; Aditya Bardia, MD; Genevieve Boland, MD, PhD, FACS; Justin Gainor, MD; Leyre Zubiri, MD, PhD (Massachusetts General Hospital Cancer Center, Boston, MA, USA)

**Thorvardur R. Halfdanarson, MD**; Tanios Bekaii-Saab, MD; Aakash Desai, MD, MPH; Zhuoer Xie, MD, MS (Mayo Clinic, AZ/FL/MN, USA)

**Ruben A. Mesa, MD, FACP**; Mark Bonnen, MD; Daruka Mahadevan, MD, PhD; Amelie G. Ramirez, DrPH, MPH; Mary Salazar, ANP; Dimpy P. Shah, MD, PhD; Pankil K. Shah, MD, MSPH (Mays Cancer Center at UT Health San Antonio MD Anderson Cancer Center, San Antonio, TX, USA)

**Gregory J. Riely, MD, PhD**; **Elizabeth V. Robilotti MD, MPH**; Rimma Belenkaya, MA, MS; John Philip, MS (Memorial Sloan Kettering Cancer Center, New York, NY, USA)

**Bryan Faller, MD** (Missouri Baptist Medical Center, St. Louis, MO, USA)

**Rana R. McKay, MD**; Archana Ajmera, MSN, ANP-BC, AOCNP; Angelo Cabal, BS; Justin A. Shaya, MD (Moores Comprehensive Cancer Center at the University of California, San Diego, La Jolla, CA, USA)

**Lisa B. Weissmann, MD,** Chinmay Jani, MD (Mount Auburn Hospital, Cambridge, MA, USA)

**Daniel G. Stover, MD**; Daniel Addison, MD; James L. Chen, MD; Margaret E. Gatti-Mays, MD; Sachin R. Jhawar, MD; Vidhya Karivedu, MBBS; Maryam B. Lustberg, MD, MPH; Joshua D. Palmer, MD; Clement Pillainayagam, MD; Sarah Wall, MD; Nicole Williams, MD (The Ohio State University Comprehensive Cancer Center, Columbus, OH, USA)

**Monika Joshi, MD, MRCP**; Harry Menon, DO, MPH; Marc A. Rovito, MD, FACP (Penn State Health/Penn State Cancer Institute/St. Joseph Cancer Center, PA, USA)

**Elizabeth A. Griffiths, MD**; Amro Elshoury, MBBCh (Roswell Park Comprehensive Cancer Center, Buffalo, NY, USA)

**Salma K. Jabbour, MD**; Mansi R. Shah, MD (Rutgers Cancer Institute of New Jersey at Rutgers Biomedical and Health Sciences, New Brunswick, NJ, USA)

**Babar Bashir, MD, MS**; Christopher McNair, PhD; Sana Z. Mahmood, BA, BS; Vasil Mico, BS; Chaim Miller, BA; Andrea Verghese Rivera, MD (Sidney Kimmel Cancer Center at Thomas Jefferson University, Philadelphia, PA, USA)

**Sumit A. Shah, MD, MPH**; Elwyn C. Cabebe, MD; Michael J. Glover, MD; Alokkumar Jha, PhD; Lidia Schapira, MD, FASCO; Julie Tsu-Yu Wu, MD, PhD (Stanford Cancer Institute at Stanford University, Palo Alto, CA, USA)

**Suki Subbiah, MD** (Stanley S. Scott Cancer Center at LSU Health Sciences Center, New Orleans, LA, USA)

**Daniel B. Flora, MD, PharmD**; Goetz Kloecker, MD; Barbara B. Logan, MS (St. Elizabeth Healthcare, Edgewood, KY, USA)

**Gilberto de Lima Lopes Jr., MD, MBA, FAMS, FASCO** (Sylvester Comprehensive Cancer Center at the University of Miami Miller School of Medicine, Miami, FL, USA)

**Karen Russell, MD, FACP**; Brittany Stith, RN, BSN, OCN, CCRP (Tallahassee Memorial Healthcare, Tallahassee, FL, USA)

**Natasha Edwin, MD**; Melissa Smits, APC (ThedaCare Cancer Care, Appleton, WI, USA)

**David Chism, MD**; Susie Owenby, RN, CCRP (Thompson Cancer Survival Center, Knoxville, TN, USA)

**Deborah B. Doroshow, MD, PhD**; Matthew D. Galsky, MD; Huili Zhu, MD (Tisch Cancer Institute at the Icahn School of Medicine at Mount Sinai, New York, NY, USA)

**Julie C. Fu, MD;** Alyson Fazio, APRN-BC (Tufts Medical Center Cancer Center, Boston and Stoneham, MA, USA)

**Jonathan Riess, MD, MS**, Kanishka G. Patel, MD (UC Davis Comprehensive Cancer Center at the University of California at Davis, CA, USA)

**Vadim S. Koshkin, MD**; Daniel H. Kwon, MD (UCSF Helen Diller Family Comprehensive Cancer Center at the University of California at San Francisco, CA, USA)

**Samuel M. Rubinstein, MD**; **William A. Wood, MD, MPH**; Jessica Yasmine Islam, PhD, MPH; Vaibhav Kumar, MD (UNC Lineberger Comprehensive Cancer Center, Chapel Hill, NC, USA)

**Trisha M. Wise-Draper, MD, PhD**; Syed Ahmad, MD; Punita Grover, MD; Shuchi Gulati, MD; Jordan Kharofa, MD; Michelle Marcum, MS; Cathleen Park, MD (University of Cincinnati Cancer Center, Cincinnati, OH, USA)

**Daniel W. Bowles, MD**; Christoper L. Geiger, MD (University of Colorado Cancer Center, Aurora, CO, USA)

**Merry-Jennifer Markham, MD, FACP, FASCO**; Rohit Bishnoi, MD; Chintan Shah, MD (University of Florida Health Cancer Center, Gainesville, FL, USA)

**Jared D. Acoba, MD**; Young Soo Rho, MD, CM (University of Hawai'i Cancer Center, Honolulu, HI, USA)

**Lawrence E. Feldman, MD**; **Kent F. Hoskins, MD**; Gerald Gantt Jr., MD; Mahir Khan, MD; Ryan H. Nguyen, DO; Mary Pasquinelli, APN, DNP; Candice Schwartz, MD; Neeta K. Venepalli, MD, MBA (University of Illinois Hospital & Health Sciences System, Chicago, IL, USA)

**Praveen Vikas, MD** (University of Iowa Holden Comprehensive Cancer Center, Iowa City, IA, USA)

**Elizabeth Wulff-Burchfield, MD;** Anup Kasi MD, MPH (The University of Kansas Cancer Center, Kansas City, KS, USA)

**Christopher R. Friese, PhD, RN, AOCN, FAAN**; Leslie A. Fecher, MD (University of Michigan Rogel Cancer Center, Ann Arbor, MI, USA)

**Blanche H. Mavromatis, MD;** Ragneel Bijjula, MD; Qamar U. Zaman, MD (UPMC Western Maryland, Cumberland, MD, USA)

**Jeremy L. Warner, MD, MS, FAMIA, FASCO**; Alex Cheng, PhD; Elizabeth J. Davis, MD; Kyle T. Enriquez, MSc BS; Benjamin French, PhD; Erin A. Gillaspie, MD, MPH; Daniel Hausrath, MD; Cassandra Hennessy, MS; Chih-Yuan Hsu, PhD; Douglas B. Johnson, MD, MSCI; Xuanyi Li, BA; Sanjay Mishra, MS, PhD; Sonya A. Reid, MD, MPH; Brian I. Rini, MD, FACP, FASCO; Yu Shyr, PhD; David A. Slosky, MD; Carmen C. Solorzano, MD, FACS; Tianyi Sun, MS; Matthew D. Tucker, MD; Karen Vega-Luna; Lucy L. Wang, BA (Vanderbilt-Ingram Cancer Center at Vanderbilt University Medical Center, Nashville, TN, USA)

**Matthew Puc, MD**; Theresa M. Carducci, MSN, RN, CCRP; Karen J. Goldsmith, BSN, RN; Susan Van Loon, RN, CTR, CCRP (Virtua Health, Marlton, NJ, USA)

**Robert L. Rice, MD, PhD** (WellSpan Health, York, PA, USA)

**Wilhelmina D. Cabalona, MD;** Christine Pilar, BS, CCRC, ACRP-PM (Wentworth-Douglass Hospital, Dover, NH, USA)

**Prakash Peddi, MD**; **Lane R. Rosen, MD**; Briana Barrow McCollough, BSc, CCRC (Willis-Knighton Cancer Center, Shreveport, LA, USA)

**Mehmet A. Bilen, MD**; Deepak Ravindranathan, MD, MS (Winship Cancer Institute of Emory University, Atlanta, GA, USA)

**Navid Hafez, MD, MPH**; Roy Herbst, MD, PhD; Patricia LoRusso, DO, PhD; Tyler Masters, MS; Catherine Stratton, BA (Yale Cancer Center at Yale University School of Medicine, New Haven, CT, USA)

**Data Dictionary used for this analysis**.
The full CCC19 data dictionary and R code to generate the derived variables is publicly available on GitHub: https://github.com/covidncancer/CCC19_dictionary

| Outcome description | Outcome variable name | Outcome values |
|---|---|---|
| 30-day all-cause mortality (primary outcome measure) | der_dead30 | 0 = No; 1 = Yes; 99 = Unknown |
| Days from SARS-CoV-2 diagnosis to death | der_days_to_death_combined | Days (integer) |
| Median f/u time | der_median_fu | Days (integer) |

| Covariate description | Variable name | Possible covariate values |
|---|---|---|
| Receipt of convalescent plasma (primary stratification variable) | der_plasma | 0 = No; 1 = Yes; 99 = Unknown |
| Age | der_age_cat | • 18-39 years<br>• 40-59 years<br>• 60-69 years<br>• 70-79 years<br>• 80+ years |
| Sex | der_sex | Male; Female |
| Race/ethnicity | der_race | • Non-Hispanic White; Hispanic; Non-Hispanic Black; Other |
| Smoking status | der_smoking2 | • Never; Current or Former; Unknown |
| Obesity | der_obesity | 0 = No; 1 = Yes; 99 = Unknown |
| Diabetes mellitus | der_dm2 | 0 = No; 1 = Yes; 99 = Unknown |
| Hypertension | der_htn | 0 = No; 1 = Yes; 99 = Unknown |
| Renal comorbidities | der_renal | 0 = No; 1 = Yes; 99 = Unknown |
| Pulmonary comorbidities | der_pulm | 0 = No; 1 = Yes; 99 = Unknown |
| ECOG performance status | der_ecogcat2 | 0; 1; 2+; Unknown |
| Method of COVID-19 diagnosis | covid_19_diagnosis | 1 = Suspected based on symptoms<br>11 = Suspected based on contact with confirmed case<br>2 = Suspected based on CXR findings<br>3 = Suspected based on CT scan findings<br>4 = Laboratory-confirmed<br>99 = Unknown |
| Baseline COVID-19 severity | severity_of_covid_19_v2 | 1 = Mild (no hospitalization required)<br>2 = Moderate (hospitalization indicated)<br>3 = Severe (ICU admission indicated)<br>99 = Unknown |
| Hospitalization (ever/never) | der_hosp | 0 = No; 1 = Yes; 99 = Unknown |
| ICU admission (ever/never) | der_ICU | 0 = No; 1 = Yes; 99 = Unknown |
| Mechanical ventilation (ever/never) | der_mv | 0 = No; 1 = Yes; 99 = Unknown |
| Hydroxychloroquine given during COVID-19 illness | der_hcq | 0 = No; 1 = Yes; 99 = Unknown |
| Remdesivir given during COVID-19 illness | der_rem | 0 = No; 1 = Yes; 99 = Unknown |
| Corticosteroids given during COVID-19 illness | der_steroids_c19 | 0 = No; 1 = Yes; 99 = Unknown |
| Tocilizumab given during COVID-19 illness | der_toci | 0 = No; 1 = Yes; 99 = Unknown |
| Absolute lymphocyte count | der_alc | • Normal<br>• High<br>• Low |

| | | |
|---|---|---|
| | | • Not drawn/Not available<br>• Unknown |
| Lymphoid malignancy (primary and/or secondary) | der_Lymph | 0 = No; 1 = Yes |
| Myeloid malignancy (primary and/or secondary) | der_Myeloid | 0 = No; 1 = Yes |
| Chronic lymphocytic leukemia | der_CLL | 0 = No; 1 = Yes |
| Plasma cell neoplasm | der_PCDs | 0 = No; 1 = Yes |
| Cancer status | der_cancer_status | • Remission/NED<br>• Active, progressing<br>• Active, stable/responding<br>• Unknown |
| Timing of anti-cancer treatment | der_cancer_tx_timing | 0 = More than 3 months prior to COVID-19<br>1 = Less than 2 weeks prior to COVID-19<br>2 = 2-4 weeks prior to COVID-19<br>3 = 1-3 months prior to COVID-19<br>88 = Never or starting after COVID-19 diagnosis<br>99 = Unknown |
| Cytotoxic chemotherapy within 3 months of COVID-19 diagnosis | der_any_cyto | 0 = No; 1 = Yes; 99 = Unknown |
| Immunotherapy within 3 months of COVID-19 diagnosis | der_any_immuno | 0 = No; 1 = Yes; 99 = Unknown |
| Targeted therapy within 3 months of COVID-19 diagnosis | der_any_targeted | 0 = No; 1 = Yes; 99 = Unknown |
| Anti-CD20 antibody cancer treatment within 3 months of COVID-19 | der_cd20 | 0 = No; 1 = Yes; 99 = Unknown |
| BTK inhibitor cancer treatment within 3 months of COVID-19 | der_btki | 0 = No; 1 = Yes; 99 = Unknown |
| Region of patient residence | der_region_v2 | Non-US; Other; Undesignated US; US Midwest; US Northeast; US South; US West |
| Dummy variable corresponding to trimester of diagnosis, for case-control matching purposes | dummy_trimester_dx | x1, x2, x3 – the mapping of these variables to actual timed events is masked as required by the collaborator agreement. |

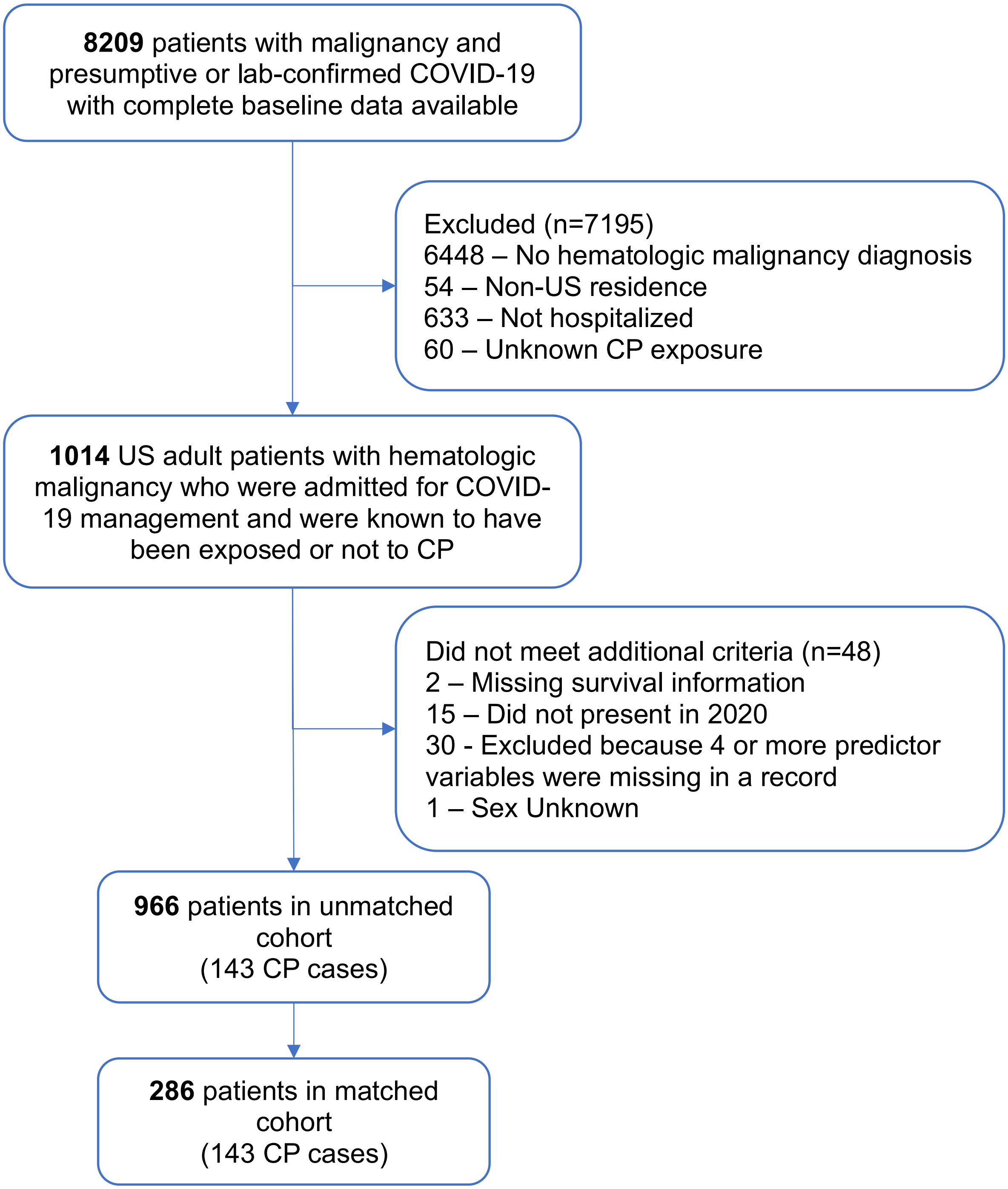

**Supplemental Figure S1: CONSORT Diagram**.

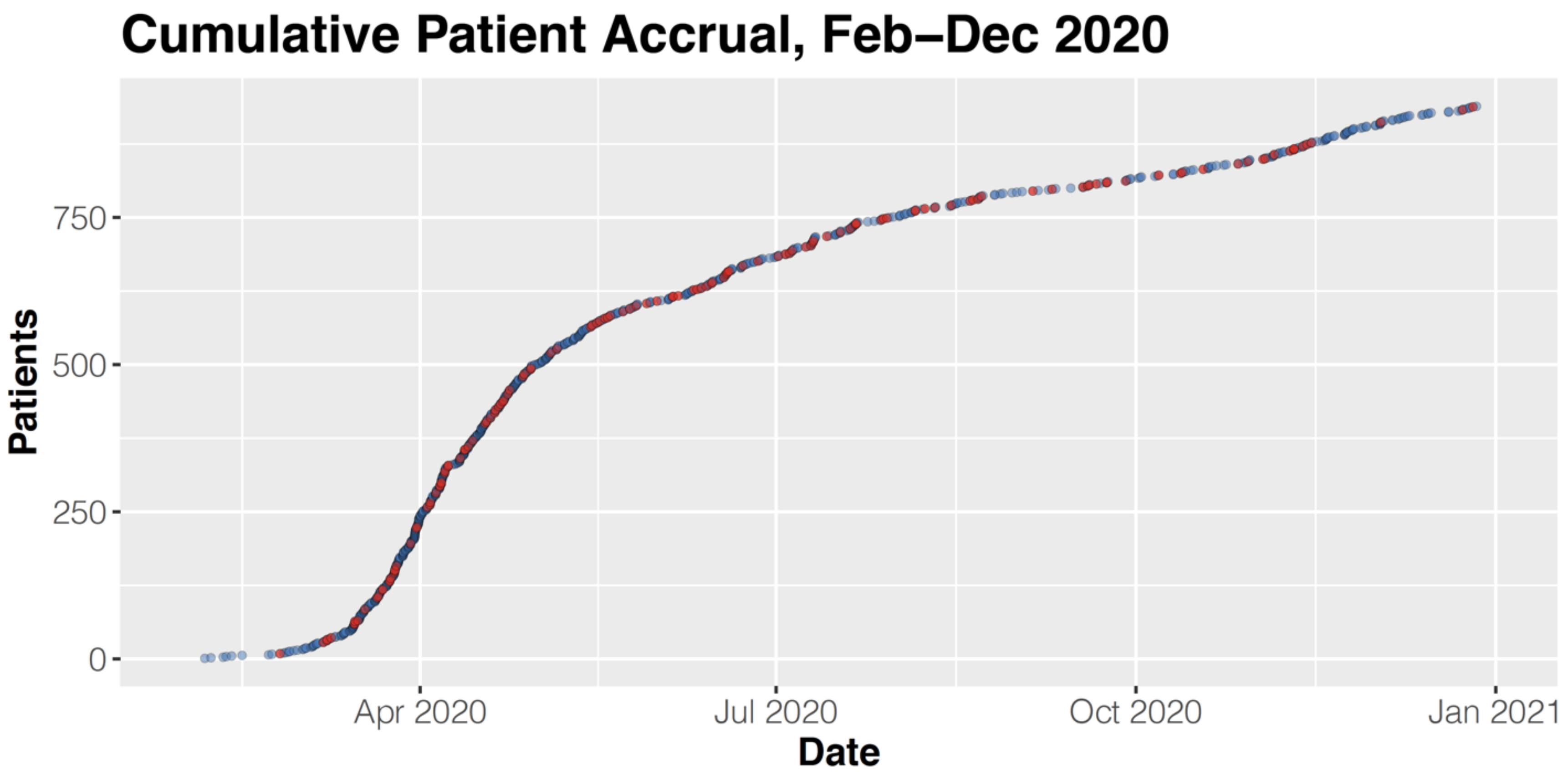

**Supplemental Figure S2: Cumulative Patient Accrual.** Red points represent patients who were treated with convalescent plasma; blue points represent patients who were not. The rate of accrual was highest in spring of 2020, with a continued near-linear growth in cases thereafter.

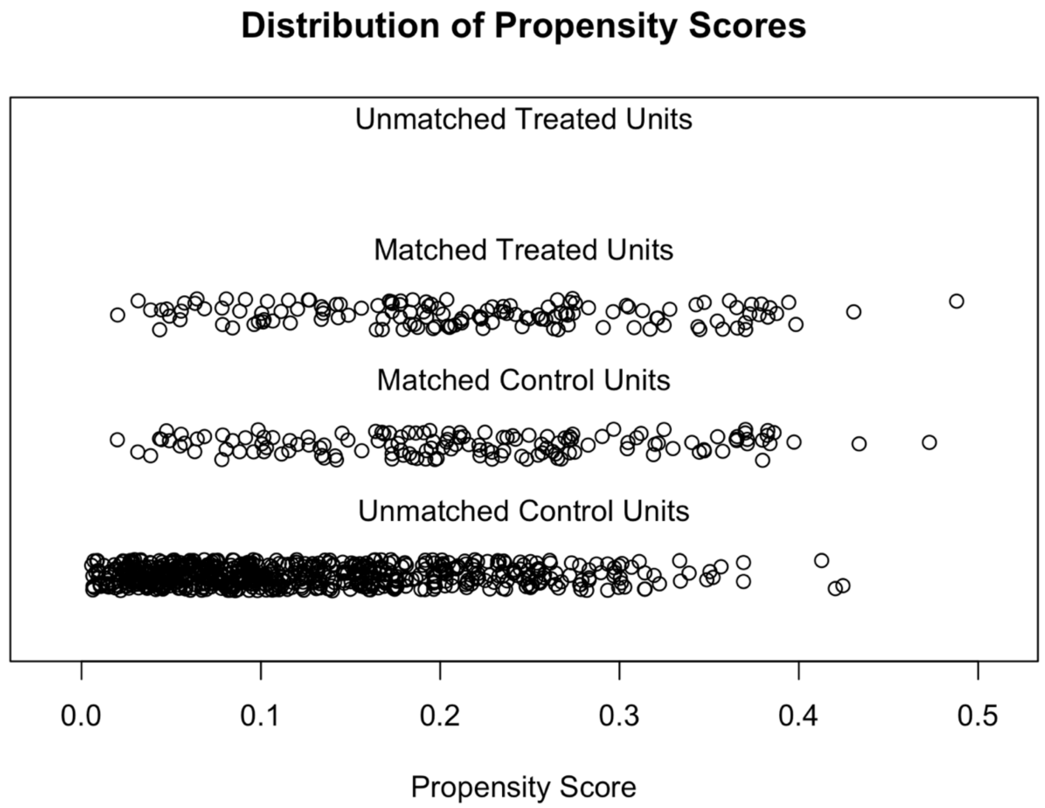

**Supplemental Figure S3: Distribution of Propensity Scores**

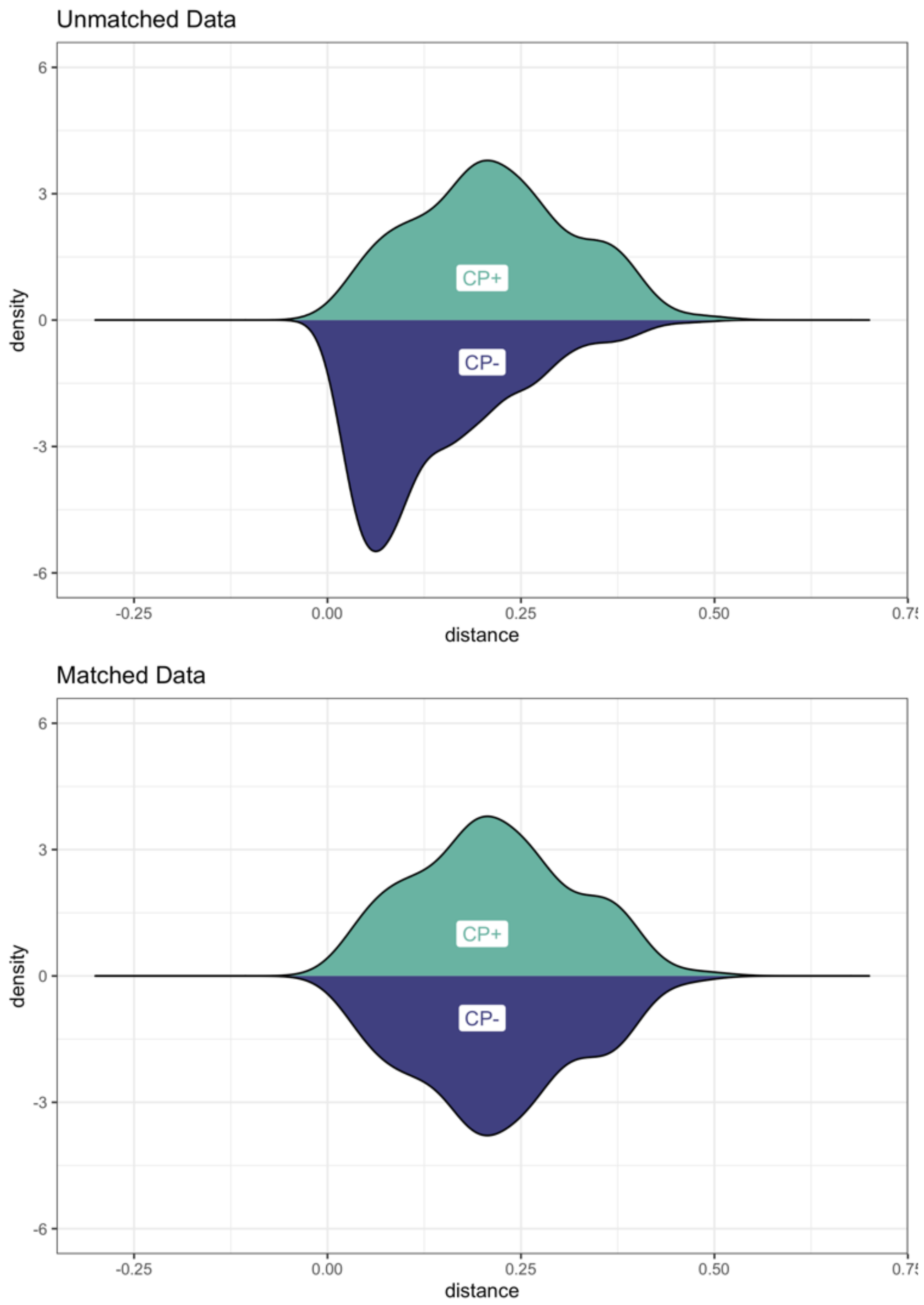

**Supplemental Figure S4: Density Graph of the Propensity Scores Before and After Matching**

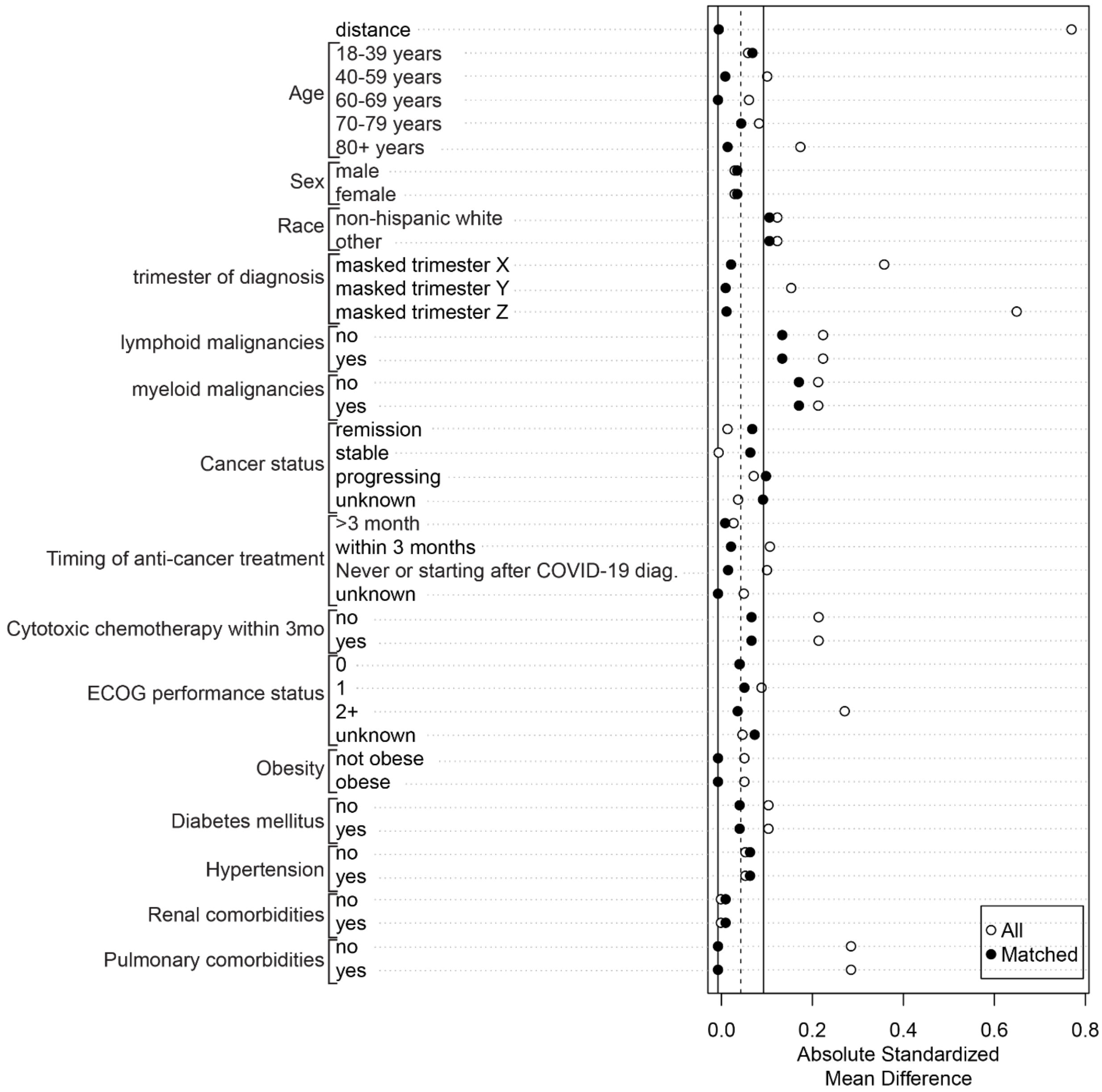

**Supplemental Figure S5: Covariate Balance.**

Distances closer to zero represent a closer match between the cases and controls; overall distance is shown at the top of the graphic.

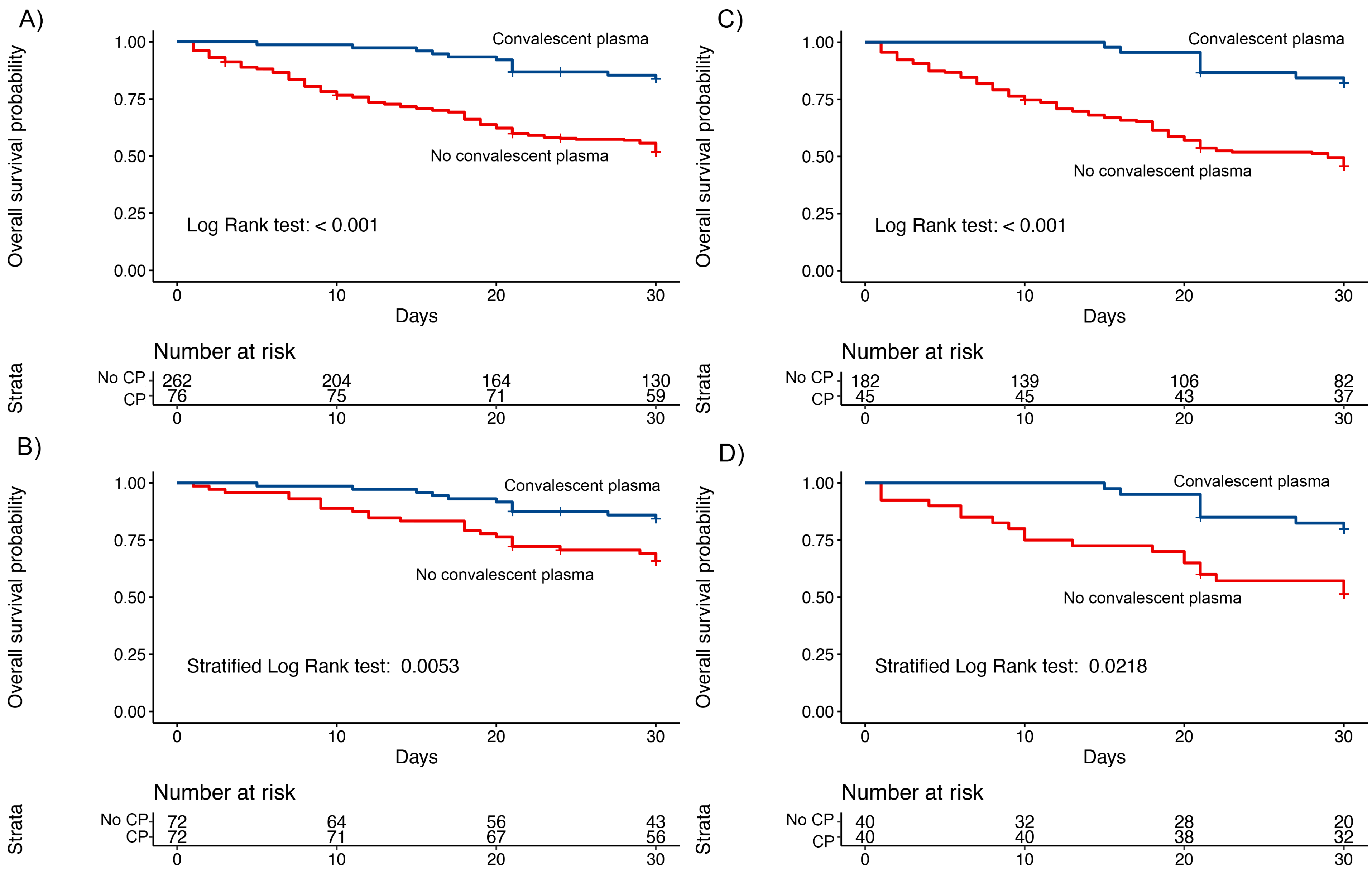

**Supplemental Figure S6: Exploratory Subgroup Analyses**. A-B) Patients admitted to the ICU: A) overall subpopulation and B) propensity-score matched population. C-D) Patients who required mechanical ventilation: C) overall subpopulation and D) propensity-score matched population.
